# Investigation of the genetic aetiology of Lewy body diseases with and without dementia

**DOI:** 10.1101/2023.10.17.23297157

**Authors:** Lesley Wu, Raquel Real, Alejandro Martinez, Ruth Chia, Michael A Lawton, Maryam Shoai, Catherine Bresner, Leon Hubbard, Cornelis Blauwendraat, Andrew B Singleton, Mina Ryten, Sonja W. Scholz, Bryan J Traynor, Nigel Williams, Michele T M Hu, Yoav Ben-Shlomo, Donald G Grosset, John Hardy, Huw R Morris, International LBD Genomic Consortium

## Abstract

Up to 80% of Parkinson’s disease patients develop dementia, but time to dementia varies widely from motor symptom onset. Dementia with Lewy bodies presents with clinical features similar to Parkinson’s disease dementia, but cognitive impairment precedes or coincides with motor onset. It remains controversial whether dementia with Lewy bodies and Parkinson’s disease dementia are distinct conditions or represent part of a disease spectrum. The biological mechanisms underlying disease heterogeneity, in particular the development of dementia, remain poorly understood, but will likely be key to understanding disease pathways and ultimately therapy development. Previous genome-wide association studies in Parkinson’s disease and dementia with Lewy bodies/Parkinson’s disease dementia have identified risk loci differentiating patients from controls. We collated data for 7,804 patients of European ancestry from Tracking Parkinson’s (PRoBaND), The Oxford Discovery Cohort, and AMP-PD. We conducted a discrete phenotype genome-wide association studies comparing Lewy body diseases with and without dementia to decode disease heterogeneity by investigating the genetic drivers of dementia in Lewy body diseases. We found that risk alleles rs429358 tagging *APOEe4* and rs7668531 near the *MMRN1* and SNCA-AS1 genes, increase the odds of developing dementia and that an intronic variant rs17442721 tagging *LRRK2* G2019S, on chromosome 12 is protective against dementia. These results should be validated in autopsy confirmed cases in future studies.

## Introduction

Parkinson’s Disease (PD), PD dementia (PDD) and Dementia with Lewy bodies (DLB), which we describe here jointly as Lewy body diseases (LBD), are characterised pathologically by alpha-synuclein aggregates forming Lewy bodies and Lewy neurites [1]. PD is a common degenerative movement disorder presenting with tremor, rigidity and bradykinesia. Non-motor features, including cognitive impairment and dementia, develop with disease progression in PD. Approximately 24% of PD patients have mild cognitive impairment at the time of diagnosis [2], and up to 80% of PD patients eventually progress to dementia (PDD) [3], which is associated with worse functioning, poorer quality of life, care home admission, and significant morbidity [4]. However, the time to dementia from motor symptom onset varies widely between patients. DLB is a synucleinopathy presenting with symptoms similar to PDD, including dementia, cognitive fluctuations, visual hallucinations, REM sleep behaviour disorder in conjunction with existing or latent parkinsonism [5]. Clinically, PDD and DLB are distinguished by the “1-year rule”, where PDD is diagnosed when dementia develops in the context of well-established PD more than one year after motor symptom onset, while a diagnosis of DLB is given when cognitive impairment precedes or coincides with motor impairment. PDD is distinguished from DLB by the temporal sequence of symptoms.

Neuropathologically, PD usually differs from PDD/DLB in the extent of Lewy body pathology in the brain, as inclusions are limited to the limbic system or brainstem in PD without dementia. However, the pathological delineation of PDD from DLB is extremely difficult as no hallmark features distinguish the two [6]. Both are characterised by Lewy bodies in cortical areas and a high frequency of Alzheimer’s disease (AD) co-pathology. Indeed, about 50% of PDD patients have beta-amyloid plaques and neurofibrillary tangles at post-mortem, which may be a better predictor of dementia than the extent of cortical alpha-synuclein pathology [7]. The majority of DLB brains also fulfil criteria for a secondary diagnosis of AD [8]. The separation of PDD and DLB as discrete clinical and pathological entities is controversial.

LBDs are primarily sporadic. Case-control genome-wide association studies (GWAS) in the past decade have identified 90 common variant risk loci associated with PD [9] and 5 risk loci associated with DLB [10]. Variation in several genes, including *GBA1*, *TMEM175* and *SNCA,* confer risk for both diseases, suggesting overlapping pathogenesis and underlying biological dysfunction. Strikingly, *TMEM175* and *SNCA* also modulate age at onset in PD [11]. On the other hand, there are distinct loci for DLB compared to PD encompassing different genes (e.g., *APOE* and *BIN1* for DLB), and in some cases distinct association signals at the same locus. In a study using targeted high-throughput sequencing, two distinct regions of the *SNCA* gene, at the 3’ and 5’ end, were found to be differentially associated with PD and DLB risk, respectively [12]. While the consequences of these distinct signals remain to be clarified, it has been hypothesised that these distinct association signals could relate to the control of gene expression in different brain regions, leading to different phenotypes [13]. Genome-wide survival analysis of PD identified *RIMS2* [14] and *LRP1B* [15] as common risk loci for progression from PD to PDD; however, they do not seem to be relevant to DLB.

Heterozygous mutations in *GBA1* are among the strongest genetic risk factors for PD and DLB [16,17]*. GBA1* encodes glucocerebrosidase, a lysosomal enzyme involved in the metabolism of glycosphingolipid. A meta-analysis of PD patients showed that *GBA1* mutations are associated with a 2.4 -fold increase in the incidence of cognitive impairment [18]. Moreover, mutation carriers tend to have earlier disease onset [11] and shorter survival [19]. In a large multicentre study of *GBA1* mutation carriers, *GBA1* was also found to be associated with PDD, with an odds ratio of 6.48 (95% CI, 2.53 – 15.37) as well as DLB, with an odds ratio of 8.28 (95% CI, 4.78– 14.88), providing evidence that *GBA1* mutations lead to impaired cognition in synucleinopathies [20]. However, as is the case for *SNCA*, the specific variants associated with PD and DLB differ [21]. Although the role of these *GBA1* variants in pathogenesis remains unclear, studies in postmortem tissue showed that reduced lysosomal GCase is associated with alpha-synuclein aggregation, inflammation and cellular damage [21], suggesting an important role for GCase in the propagation of alpha-synuclein pathology. This could explain the spread of Lewy bodies to limbic and neocortical areas of PD patients with *GBA1* mutations.

The apolipoprotein E (*APOE*) ε4 allele, a well-known risk locus for AD, has also been identified as a strong genetic risk factor for developing PDD/DLB [10]. *APOE4* promotes amyloid-beta oligomerization and its pathological accumulation [22]. The role of *APOE4* in DLB pathogenesis is still unclear. It has been suggested that *APOE4* might be a driver of amyloid-beta deposition, which presents as a co-pathology in the majority of DLB brains [8]. However, there is some evidence showing *APOE* may contribute to cognitive decline independently of amyloid. In autopsy studies, *APOE4* was associated with dementia and diffuse LB pathology in “pure” DLB patients (i.e., with absent or low levels of amyloid) as well as PDD [23,24]. Mouse models of synucleinopathy have also demonstrated that *APOE4* exacerbated alpha-synuclein pathology in the absence of amyloid [25,26].

PD, PDD and DLB share common risk genes. However, specific risk loci within these genes may vary across the diseases, potentially leading to different phenotypes, which ultimately relate to the involvement of different cell types. Previous GWAS for PD and DLB have compared PD and DLB cases to controls. Here, in a study of almost 8000 cases, we aim to define the genetic determinants of dementia in LBD by taking a different approach. We have taken a disease classification agnostic approach by comparing all Lewy body diseases with dementia (LBD-D), including both PDD and DLB, to Parkinson’s disease cases without dementia (LBD-ND). This “case-case” approach should help identify specific variants that are associated with more extensive LB and AD pathology that contribute to cognitive impairment, rather than variants that are related to the initiation of LB pathology as compared to unaffected controls.

## Methods

### Cohort description and study design

We analysed 3 large independent cohorts: Tracking Parkinson’s (TDP, www.parkinsons.org.uk/) [27], Oxford Parkinson’s Disease Centre Discovery (OPDC, www.dpag.ox.ac.uk/opdc/) [28] and Accelerating Medicine Partnership - Parkinson’s Disease Initiative (AMP-PD v2.5, https://www.amp-pd.org/) (Supplementary Tables, Table 1). The AMP-PD dataset is enriched for patients with *LRRK2* p.G2019S. Participants were included in the present study based on their most recent clinical diagnosis or final pathological diagnosis of PD, PDD or DLB. A status of “case” for LBD-D was defined if the patient had a clinical diagnosis of DLB [5] or met the Movement Disorder Society task force PDD diagnostic criteria [29]. In detail, for PDD the criteria included: 1) scoring below the threshold for dementia on the Montreal Cognitive Assessment (MoCA score <21/30), 2) having cognitive deficits which are severe enough to interfere with activities of daily living (MDS-Unified Parkinson’s disease Rating Scale (UPDRS) part I 1.1 ≥ 2 score), 3) and the absence of severe depression defined using the MDS-UPDRS (MDS-UPDRS part I 1.3 < 4). LBD-ND were given a status of “control”. The patients did not have dementia based on the available clinical data we analysed. Patients with a change of diagnosis to a non-Lewy body disorder during the follow-up period were removed from analyses. AMP-PD is a unified cohort consisting of eight longitudinal studies with similar sample collection protocols. All studies were approved by local and multi-centre ethics committees and are in compliance with the Declaration of Helsinki. Appropriate data use agreements were approved.

**Table 1.**
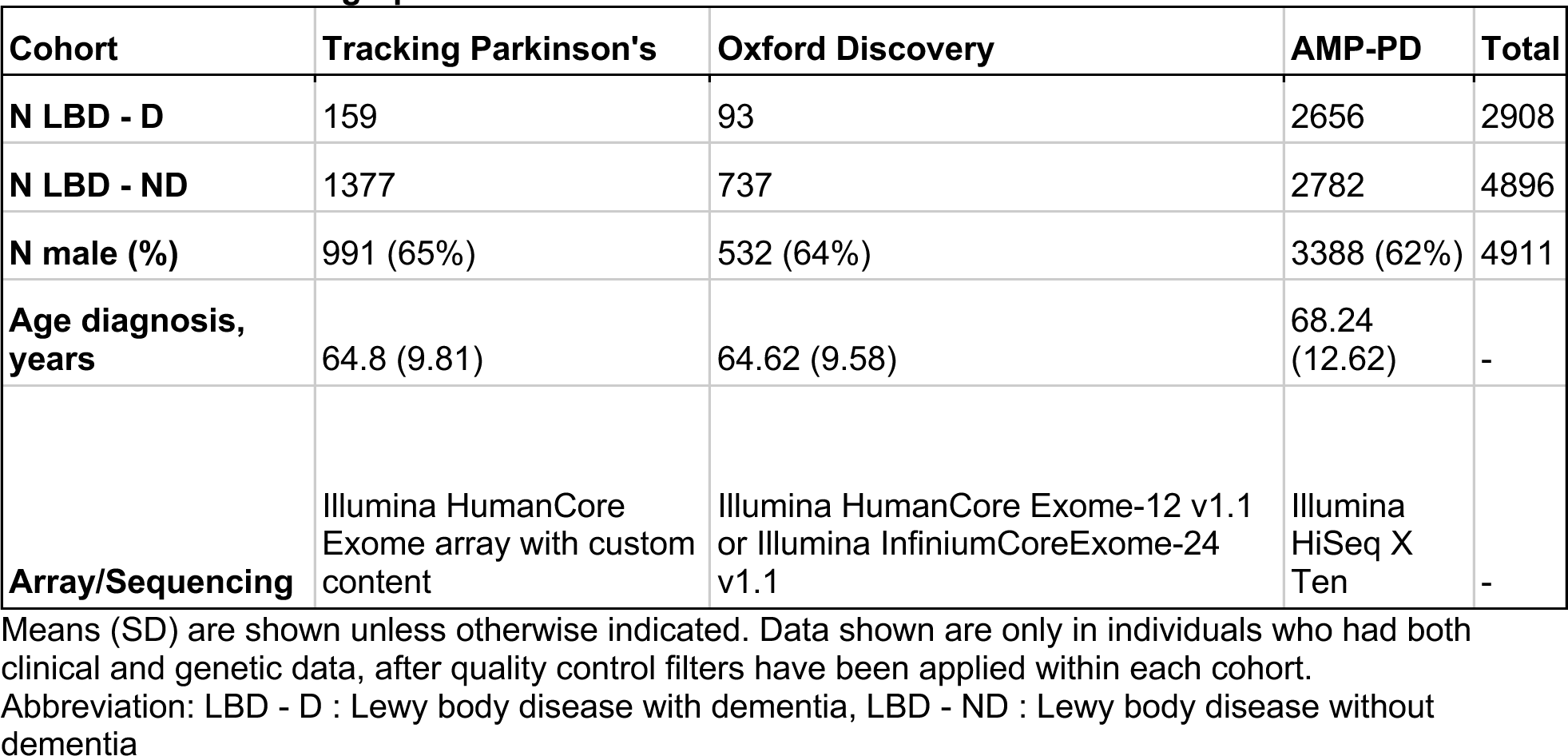
Cohort demographics.

### Genotyping and quality control

DNA was extracted from whole blood or brain tissue as detailed in the protocols of each study. TPD used the Illumina HumanCoreExome array with custom content for genotyping. OPDC generated genotype data using the Illumina HumanCoreExome-12 v1.1 and Illumina Infinium HumanCoreExome-24 v1.1 SNP arrays. Whole genome sequencing for AMP-PD samples was performed using Illumina HiSeq X Ten sequencer and data was processed against Human Genome Reference Build 38 (https://ftp.1000genomes.ebi.ac.uk/vol1/ftp/technical/reference/GRCh38_reference_genome/). Data cleaning was performed using PLINK v1.9 (RRID:SCR_001757; https://www.cog-genomics.org/plink/1.9/ [30]) and PLINK v2.0 (https://www.cog-genomics.org/plink/2.0/). For quality control at sample-level, we excluded individuals from analysis if they had a low genotyping call rate (<= 95%), excessive heterozygosity rates (> +/- 0.15 F-statistic), or mismatch between clinically reported and genetically determined sex by the X chromosome. We also excluded duplicate or related samples (kinship coefficient > 0.088). We removed individuals that were not of European ancestry by performing a principal component analysis from pruned genetic data of each cohort included in the analysis. We used Hapmap3 as the reference panel to derive ancestry groups. Individuals that deviated by more than 3 standard deviations from the mean of the first 2 principal components of the HapMap3 CEU group were removed from the analysis.

For quality control at the variant-level, we removed SNPs from analysis if they had a low genotyping rate (< 0.99%), deviated significantly from Hardy-Weinberg equilibrium (P < 1E-8), had a minor allele frequency less than 1%, and were non-autosomal (X, Y, mitochondrial chromosomes). After quality control, genetic data for TDP and OPDC were imputed separately against the TOPMed (https://imputation.biodatacatalyst.nhlbi.nih.gov/33)[31] r2 panel with Eagle v2.4 phasing on the TOPMed Imputation Server using Minimac4 [56,57]. We used the Rsq info measure of imputation accuracy to exclude variants that were not confidently imputed. We filtered out variants with an Rsq lower than 0.8. We also removed SNPs if missingness was > 5% and minor allele frequency was < 1%. The two datasets were then merged, with only shared variants retained.

### Statistical analysis for single-variant associations

Clinical data was cleaned and analysed using R v4.1.3 (RRID:SCR_001905;R Project for Statistical Computing, version 4.1.3; https://www.R-project.org/). We used logistic regression in PLINK to perform two separate genome-wide association studies for LBD-D (DLB and PDD) compared to LBD-ND (PD cases without dementia), in AMP-PD and in the merged TDP/OPDC datasets, respectively. The following covariates were incorporated in our model: age at onset (for TDP/OPDC cohorts) or age at diagnosis (for AMP-PD), sex and the first five genetic principal components. We meta-analysed the summary results for TDP/OPDC and AMP-PD using METAL (RRID:SCR_002013;http://csg.sph.umich.edu//abecasis/Metal/) [32], under a random effects model, using genomic control correction. We only included variants present in all cohorts, and with minor allele frequency variability below 15% across studies. We used Cochran’s Q to test for heterogeneity in the meta-analysis and excluded variants with p-value < 0.05 and I^2^ statistic =< 80%. We considered p-values below 5 x 10^-8^ to be genome-wide significant, and nominally significant below 5 x 10^-6^. We used Locuszoom to generate the Manhattan plot and the regional association plots (RRID:SCR_021374; http://locuszoom.org/)[33].

### Conditional analysis

In order to determine whether there were single or multiple independent signals at each genome-wide significant locus, we carried out a conditional and joint multiple-SNP analysis (COJO) on the GWAS summary statistics. We used the AMP-PD cohort as the reference panel to estimate the LD between SNPs and apply corrections to the models, as it is the largest participating cohort in the meta-analysis. COJO was performed using GCTA (v1.93.0,GCTA | Yang Lab)[34].

### Colocalization analysis

We performed a colocalization analysis to investigate whether there is a shared causal variant between the risk of dementia in LBD cases and expression quantitative trait loci (eQTLs). We used the *coloc* R package (version 5.1.0; https://cran.rproject.org/web/packages/colocr/index.html)[35] and *colochelpR* as a wrapper (version 0.99.0)[36]. Coloc is based on Bayesian statistical approach to compute a posterior probability (PP) for the following hypotheses: there is no association with either trait (H0), there is an association with the Lewy body dementia trait but not the eQTL trait (H1), there is an association with the eQTL trait but not the Lewy body dementia trait (H2), there is association with a Lewy body dementia and an eQTL variant, but the causal variants are independent (H3), and there is a shared causal variant associated with Lewy body dementia and eQTL within the analysed region (H4). *Coloc* was run using default per SNP priors p_1_=10^−4^, p_2_=10^−4^, and p12=10−5. A PPH4 > 0.80 was considered statistically significant support for colocalization. We used Cis-eQTL data from eQTLGen, which includes 31,684 individuals (https://www.eqtlgen.org/cis-eqtls.html) and compares genetic variation with blood RNA, and PsychEncode. PsychEncode includes 1,387 individuals (http://resource.psychencode.org/) and compares genetic variation with brain RNA. We extracted all the genes from +/- 1Mb of the significant hits from the GWAS and performed a colocalization analysis on each gene. Since the cis-eQTL and the GWAS summary statistics were in different builds, we converted the summary statistics of the meta-analysis from hg38 to hg19 using the liftOver tool (RRID:SCR_018160;https://genome.sph.umich.edu/wiki/LiftOver).

### Polygenic risk score

To assess whether PDD is genetically more similar to PD or DLB, as well as the overlap with the AD risk profile, we computed a polygenic risk score (PRS) on 357 PDD patients identified across the cohorts included in the GWAS. We used previously published PD, DLB and AD GWAS [9,10,37] as the reference data. After performing QC on summary statistics of the base datasets, we used PRSice-2 (version 2.3.5; RRID:SCR_017057; https://choishingwan.github.io/PRSice/) [38] to calculate PRS with the C + T method, which involves clumping SNPs and performing p value thresholding. After clumping, 1,284,510 SNPs were included to generate the PD PRS, 380,274 SNPs for the DLB PRS and 11,931 for the AD PRS. We then conducted a general linear regression adjusted for age at onset, sex and PC1 - PC5 to test if the PRS predicted the development of dementia. Results from the regression were meta-analysed in R with the meta package (RRID:SCR_019055; https://cran.r-project.org/web/packages/meta/index.html).

## Results

After QC, a total of 7,804 individuals were selected, including 2,908 LBD-D (2552 DLB, 357 PDD) and 4,896 LBD-ND. Case selection is summarised in Supplementary Figure 1 Supplementary figures). Demographic characteristics are summarised in Table 1. LBD-D patients were significantly older than LBD-ND at diagnosis (Kruskal-Wallis chi-squared = 2627, df = 1, P-value < 2.2e^-16^.

### Identification of risk loci for dementia in LBD

Using a case-case GWAS approach comparing patients with LBD-D and LBD-ND, we analysed 6 226 081 SNPs and identified three genome-wide significant loci (Figure 1, Table 2)

**Figure 1.**
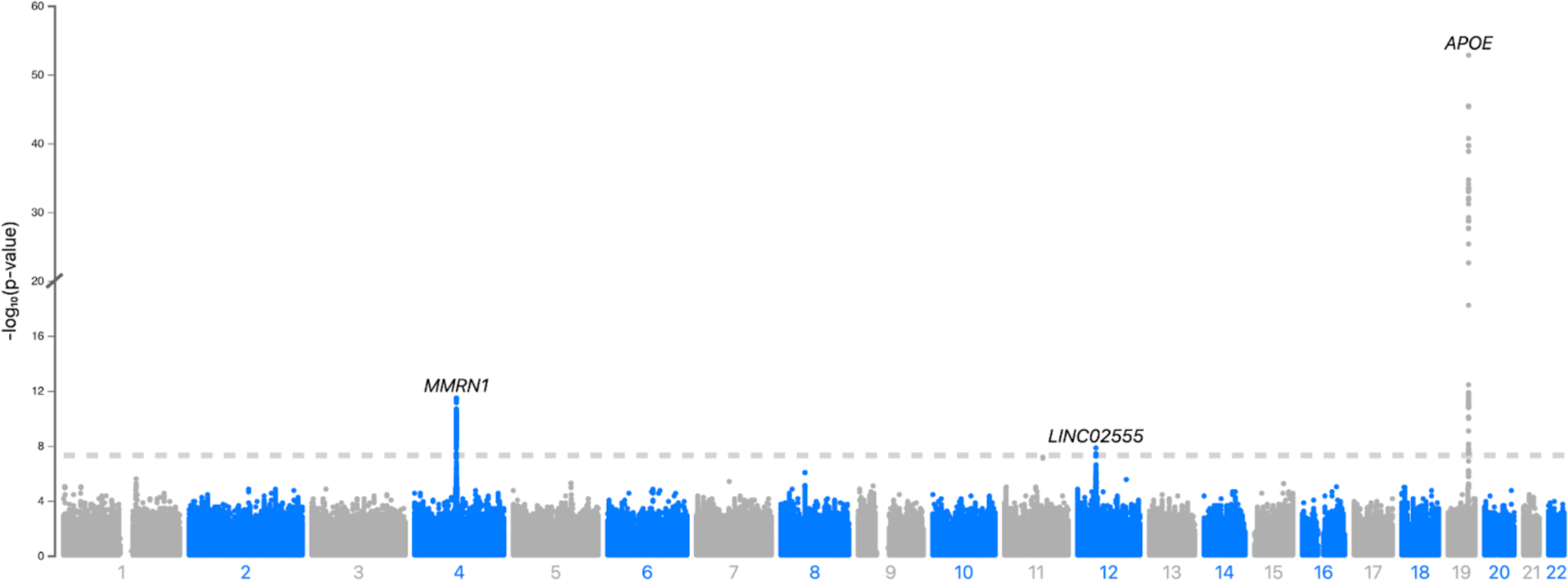
Manhattan plot of LBD - D vs LBD – ND. A Manhattan plot representing the results of the GWAS comparing LBD with dementia (LBD-D) to LBD without dementia (LBD-ND), highlight genome-wide significant SNPs on chromosome 4, 12 and 19. Negative logarithm p-value is represented on the Y axis while chromosome position is represented on the X axis. The dotted line indicates genome-wide significant threshold (5×10-8)

**Table 2.**
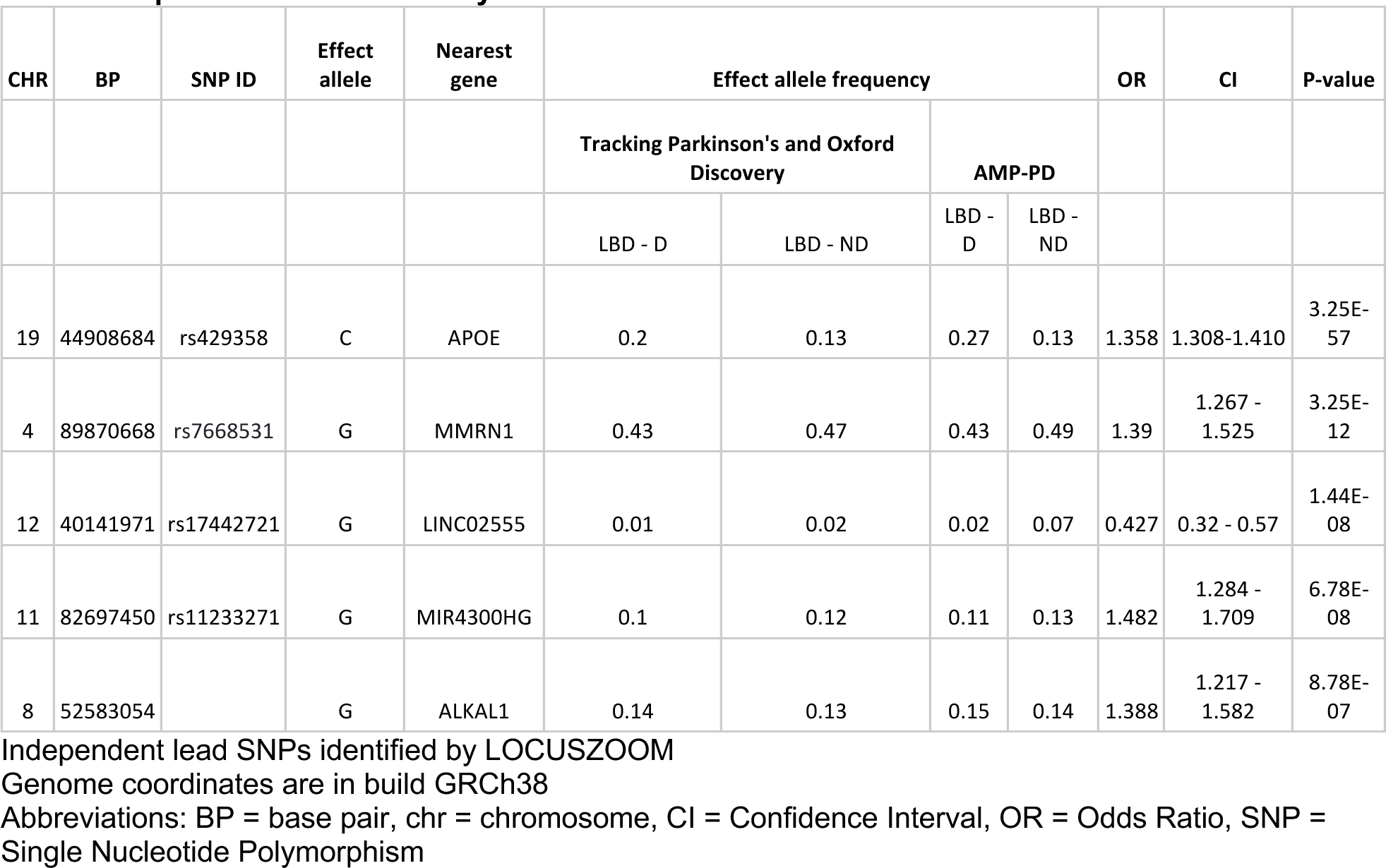
Top SNPs from meta-analysis.

The lead SNP was rs429358 in the *APOE* gene on chromosome 19 (OR = 1.36, 95% CI = 1.31 - 1.41, P = 3.25e-57). *APOE* encodes apolipoprotein E, a known genetic factor for AD and DLB. It has also been identified as a risk factor for dementia in PD [39,40]. Conditional analysis on the lead SNP detected a secondary signal at the *APOE* locus, at 19:32848205.

The second genome-wide significant SNP was rs7668531 between the *MMRN1* gene and the *SNCA* gene (OR = 1.39, 95% CI = 1.267 - 1.525, P = 3.25E-12), located 170,323 kb downstream of the *SNCA* gene. This SNP is close to and in linkage disequilibrium with rs7680557 (D’ = 0.9959, r^2^ = 0.9196), which is associated with dementia as identified in the most recent LBD case-control GWAS [10]. The rs7668531 signal is no longer genome-wide significant when we condition on the top rs7680557 in our dataset, which suggests that rs7668531 is not independent and most likely tags *SNCA-AS1*.

The third genome-wide significant SNP was rs17442721 in the noncoding RNA *LINC02555*, which was protective against developing dementia (OR = 0.43, 95% CI = 0.32 - 0.57, P = 1.44e 08). *LINC02555* is potentially a regulatory locus for *LRRK2* expression in specific cell types [41] and may mediate PSP survival [42]. However, this SNP is in LD with *LRRK2 G2019S* (rs34637584, r2 = 0.54, D’ = 0.97). To confirm whether rs17442721 is independent of *LRRK2 G2019S*, we performed a conditional analysis. Results show that rs17442721 is no longer genome-wide significant after conditioning on the G2019S variant, confirming that it tags *LRRK2* G2019S, and there is no difference in dementia related to this SNP when the data is stratified by G2019S status. In this dataset, the rate of dementia in *LRRK2* G2019S carriers is 5% as compared to 39% in the total dataset (Supplementary Tables). rs17442721 was not in linkage disequilibrium with PSP progression variant rs2242367 (r2<0.05).

Rs11233271 on chromosome 11 near the MIR4300HG gene approached genome-wide significance (OR = 1.48, 95% CI = 1.28 - 1.71, P = 6.78e-08), although this will need further evaluation in future work.

Common variant GBA E326K was nominally but not genome-wide significant (OR = 2.01, 95% CI = 1.44 - 2.83, P = 2.517e-06). The PD case-control GWAS LRRK2 rs76904798 variant was also not genome-wide significant (OR = 1.02, 95% CI = 0.88 - 1.18, P = 0.7759).

### Colocalization

We performed colocalization analysis to assess the probability of a shared causal signal between dementia status and genetically determined gene expression regulation. eQTLs were obtained from eQTLGen and PsychENCODE. eQTLGen comprises gene expression derived from blood, and psychENCODE dataset comprises gene expression from bulk RNA sequencing from the frontal cortex. We found suggestive colocalization between the genome-wide significant signal on chromosome 12 and cis-eQTL data from eQTLGen (PPH4 = 0.7154) for LRRK2 and rs11233271 on chromosome 11 suggestively colocalized with FAM181B (PPH4 = 0.7009), a protein-coding gene that is expressed in the brain (Figure 2).

**Figure 2.**
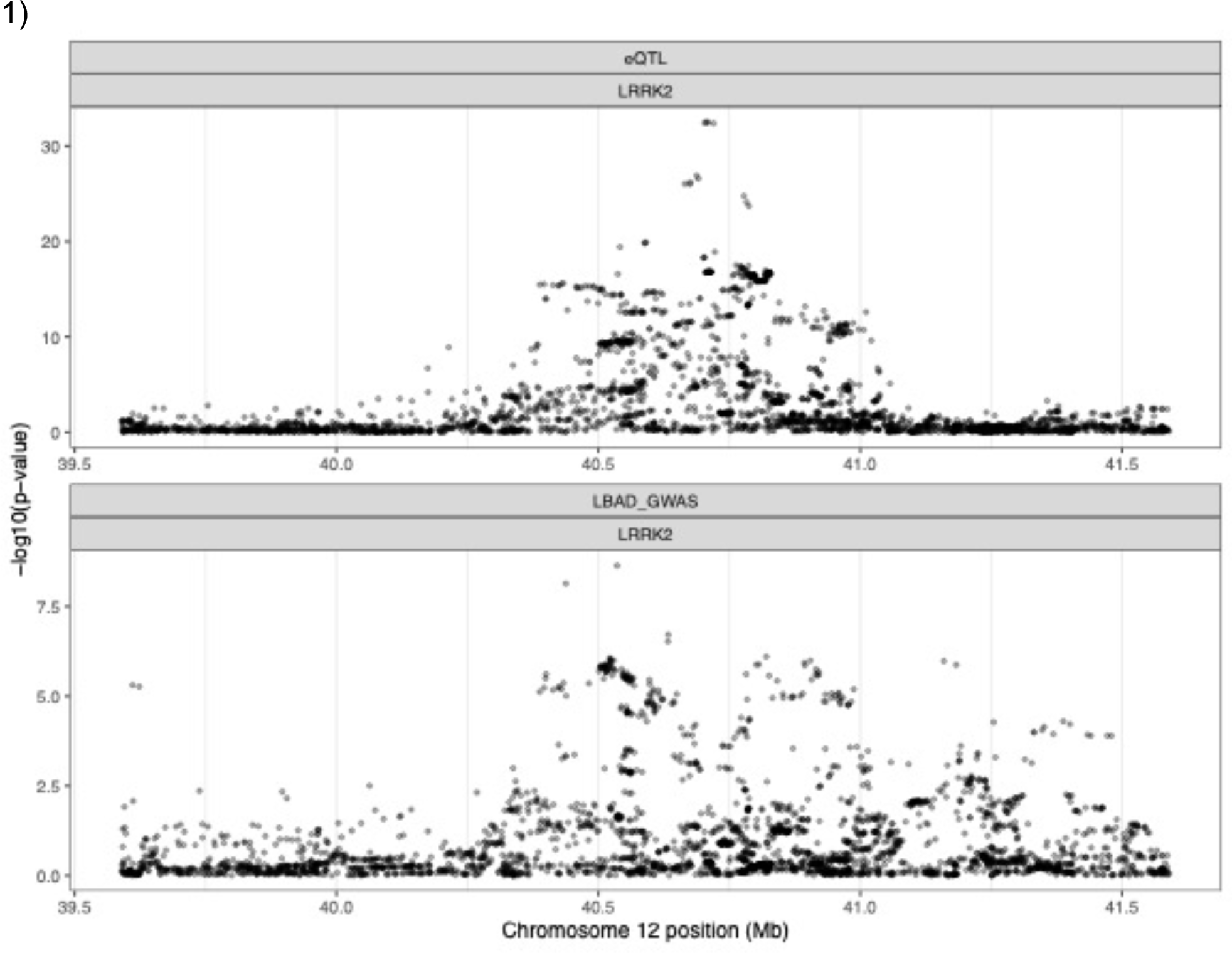
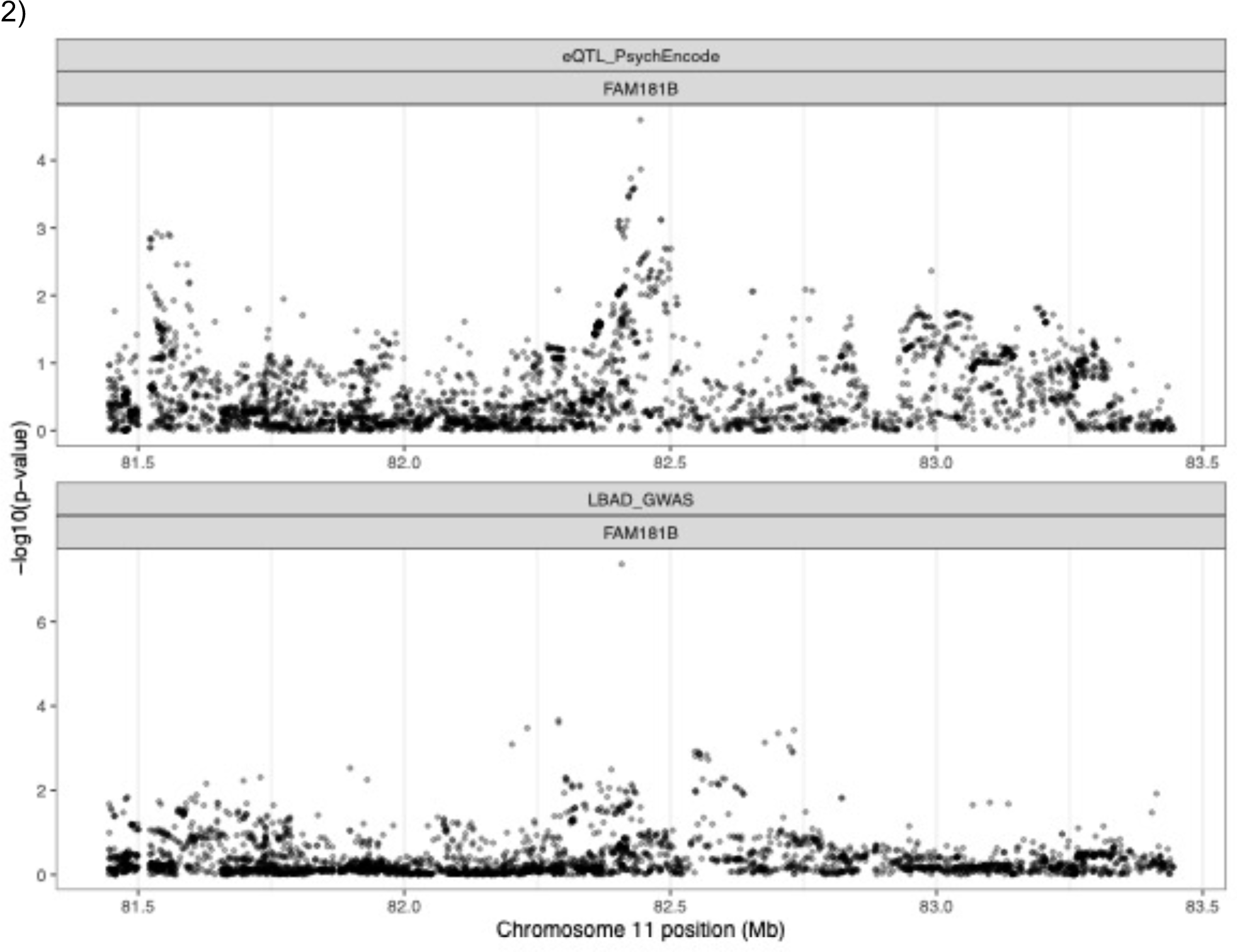
Regional association plot for eQTL and GWAS signals. Regional association plot for eQTL and 1) GWAS signal in the region close to LRRK2 (PPH4 = 0.72) and 2) in the region close to FAM181B (PPH4 = 0.70). Negative logarithm p-value is represented on the Y axis while chromosome position is represented on the X axis.

### Polygenic risk score

We applied a PD, AD and DLB polygenic risk score (PRS) derived from the most recent GWAS to PDD patients identified in each of our cohorts, as well as to PD patients for comparison. We used a general linear regression model to assess if the normalised PRS predicted dementia and meta-analysed the regressions using a random effects model. Optimised p-values PRS, based on genome-wide significant and sub-genome significant SNPs, indicated that patients with a higher PD PRS score (based on 1,284,510 SNPs) were less likely to develop dementia (OR = 0.74, 95% CI = 0.56 -0.98, p = 0.03), and the AD risk profile (based on 31000 SNPs) was not significantly different between the 2 groups (OR = 0.99, 95% CI = 0.82 - 1.20, p = 0.93). PDD was significantly associated with higher (pure) DLB PRS (OR = 2.69, 95% CI = 0.69 - 10.42, p = 0.01); however, this needs to be interpreted with caution as the confidence interval is very large (Figure 3).

**Figure 3.**
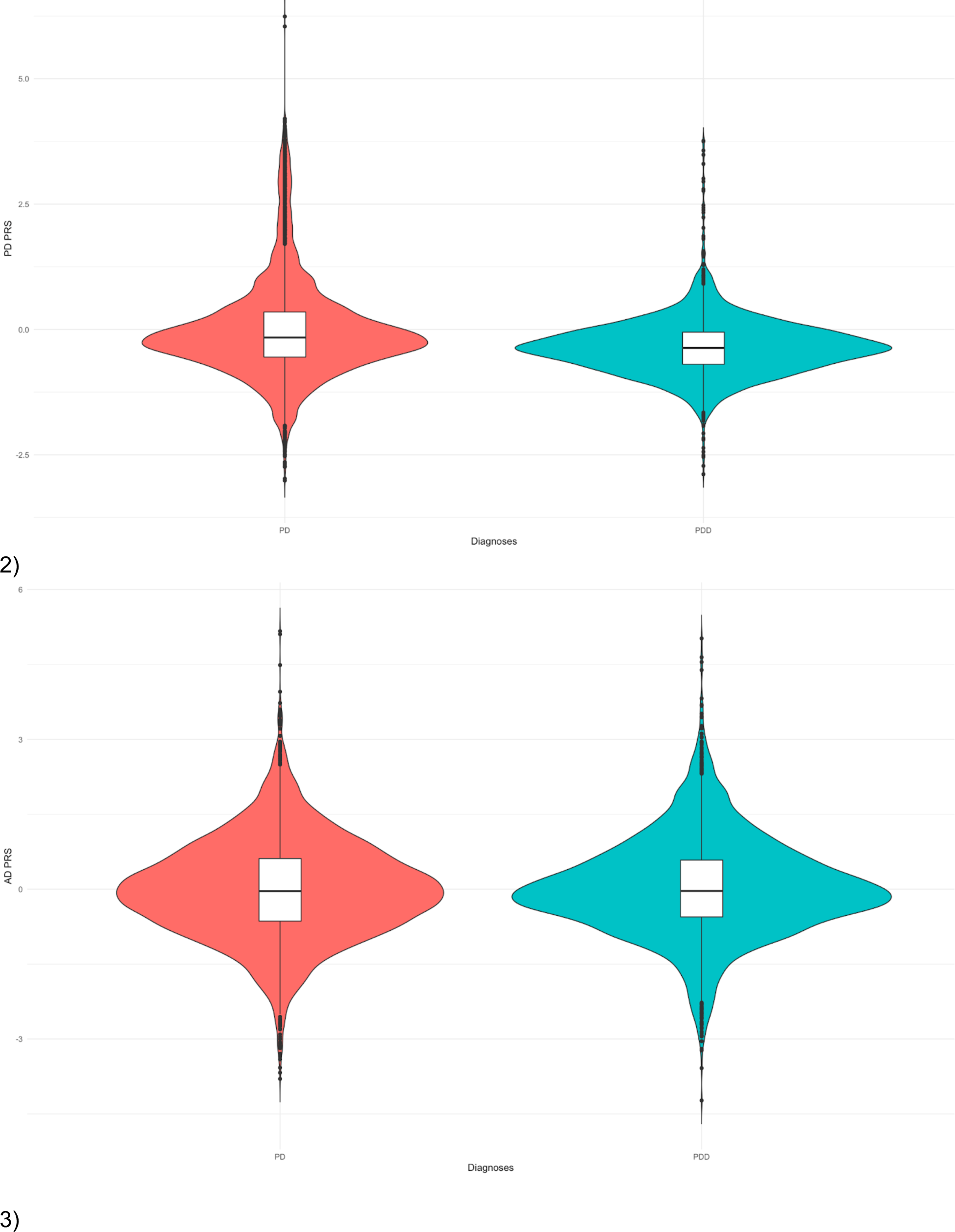
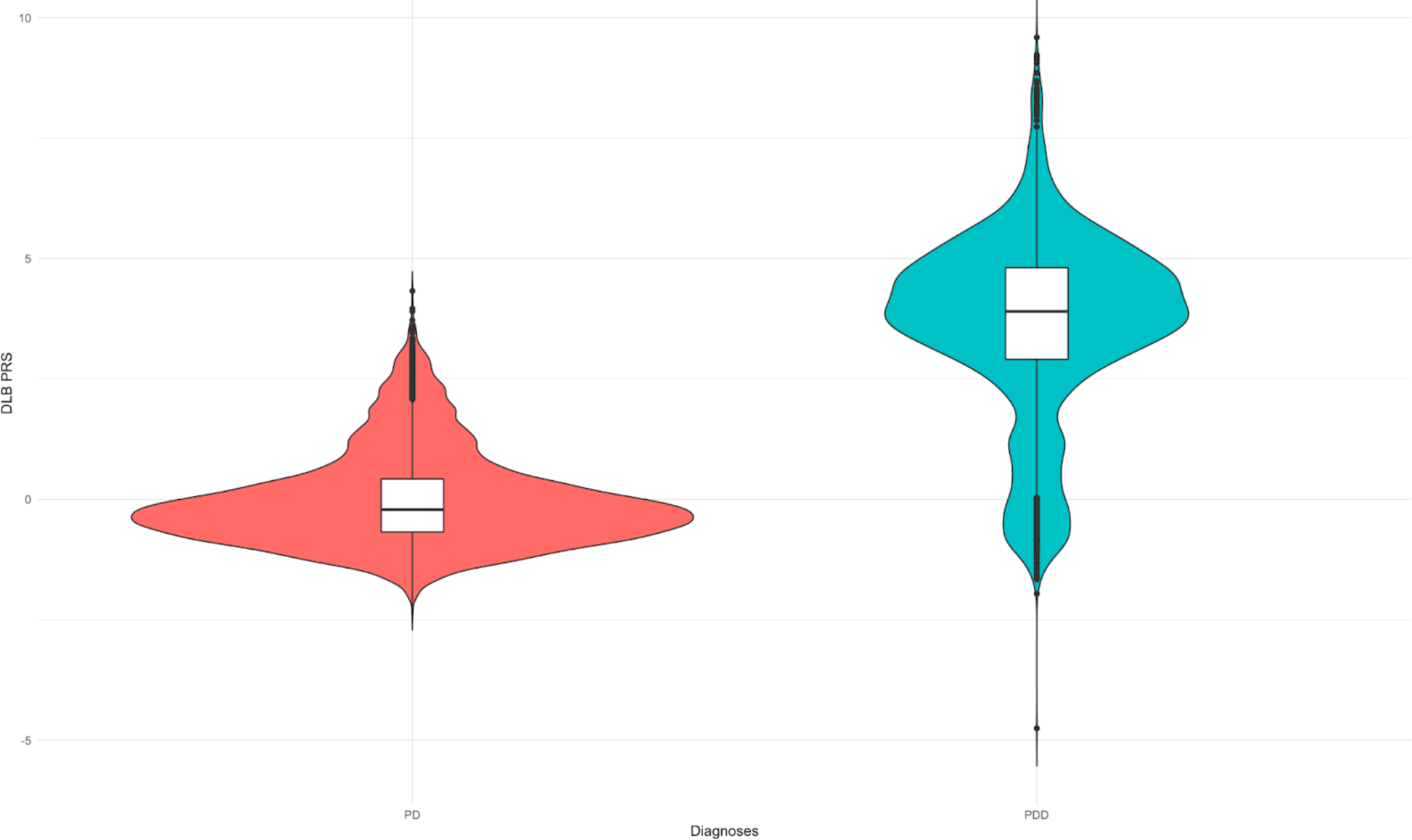
Polygenic risk score from PD, AD and DLB GWAS. Violin plot comparing z-transformed 1) Parkinson’s disease, 2) Alzheimer’s disease and 3) Dementia with Lewy body polygenic risk score distributions in PD and PDD. The centreline of the box plot represents the median and the box limits are the interquartile range. Dots correspond to outliers.

## Discussion

We have conducted a large-scale genome-wide case-case analysis to understand the genetic drivers of dementia in Lewy body diseases, by comparing LBD with and without dementia and identified three independent genome-wide significant signals in a novel case-case analysis by comparing LB cases with dementia with cases unaffected by dementia.

In line with previous studies, we showed that *APOE e4* is the strongest risk factor for dementia in LBD. Given the role of *APOE e4* in AD, this may modulate the risk of dementia via AD pathology in at least a subset of the LBD-D cases; however, previous work has been inconsistent. A substantial proportion (30-40%) of patients with PD and 50-80% of patients with DLB have co-occurring AD pathology [43]. However, it is unclear whether *APOE e4* drives dementia via AD pathology or independently. Our results indicate that the AD PRS does not drive PD dementia, suggesting that APOE e4 may drive dementia in PD cases by an AD pathology-independent mechanism. Consistent with our findings, post-mortem studies have found that *APOE e4* was associated with dementia in LBD in both “pure” LBD and those with AD co-pathology [24]. More recently, Kaivola et al. investigated in detail a subset of autopsied DLB patients which were included in the AMP-PD cohort [44]. In contrast to the previous studies, they found that APOE e4 was associated with DLB with AD pathology, but not pure DLB. Further research is needed to clarify the role of *APOE e4* in LBD. This is important as in the advent of availability of disease-modifying drugs such as lecanemab [45], some LBD-D patients might also benefit from these therapies.

We also found a SNP between MMRN1 and the 5’ end of *SNCA* at the *SNCA-AS1* locus but not at the 3’ end to be significantly associated with dementia, consistent with previous candidate gene studies [12] and GWAS [10]. *SNCA-AS1* encodes a long non-coding RNA. Cell work has demonstrated that overexpression of *SNCA-AS1* increases expression of *SNCA* mRNA [46], and so this may increase dementia risk by a direct SNCA over-expression process. Post mortem studies have found that alpha-synuclein in cortical areas is a predictor of dementia in LBD. The finding that *SNCA-AS1* is specific to LBD-D makes it an interesting potential therapeutic target. Indeed, LBD-D tends to have a much more aggressive disease course, with faster progression to mortality.

Targeting *SNCA-AS1* could therefore be a potential solution to reduce alpha-synuclein pathology in the cortex and the progression to dementia in LBD. Our study has separated the 3’ signal in SNCA, which is associated with PD risk from the 5’ signal associated with PDD. We hypothesise that the 3’ signal is important for the level of *SNCA* expression and the initiation of the PD process, particularly in subcortical areas, whereas the 5’ SNP is associated with the expression of *SNCA* in cortex [47]. Indirectly, this suggests that local *SNCA* expression is important, distinct from cell to cell spread from subcortical areas.

The third genome-wide significant signal was located near *LINC02555*, which is potentially a regulator of *LRRK2*. However, we confirmed that this SNP is tagging *LRRK2* G2019S. In the present study, we did not exclude *LRRK2* mutation carriers in the main analysis. As previously described, we have shown in this study that *LRRK2* G2019S carriers are less likely to develop dementia [48]. Moreover, *LRRK2* likely does not play a major role in DLB [49]. Our results confirm that in LBD, *LRRK2* G2019S mutation status is associated with a decreased odds of progression towards dementia.

Rs11233271 on chromosome 11, close to MIR4300HG, was nominally significant in our GWAS. This SNP may regulate the expression of *FAM181B*, a protein-coding gene involved in the development of the nervous system [50]. *FAM181B* was also associated with working memory in a gene-based study on cognitive measures in adolescence [51]. Furthermore, this locus has been associated with variation in the microbiome. Further studies are needed to investigate the role of this locus in Lewy body dementia.

Interestingly, *GBA1, BIN1 and TMEM175*, which are associated with case-control LBD GWAS [10] did not appear significant when comparing LBD-D to LBD-ND. Since *GBA1* is a known risk gene for both PD and DLB, our analysis shows that variation in *GBA1* does not distinguish between LBD-D and LBD-ND, within a study of this size. Similarly, *TMEM175* is a risk factor in both LBD with and without dementia. Therefore, it is not surprising that the signal disappears when we make a head-to-head comparison. *BIN1* encodes bridging integrator 1 and is the second strongest signal associated with AD but was not genome-wide significantly associated with LBD-D in this study (p=2.276e-05) [52]. Increased *BIN1* expression is associated with higher load of tau in the AD brain, but not amyloid [53]. While some studies found tau load to be a correlate of dementia in PD and DLB, other studies have not. Autopsy studies have found tau to colocalize with alpha-syn in Lewy bodies in both PD and DLB [54]. A small autopsy study in LRRK*2* carriers found that 100% of the brains had tau pathology [55]. Therefore, it is possible that LBD risk genes associated with tau pathology are not good candidates to distinguish LBD-D from LBD-ND. *RIMS2* was identified as a progression locus in a genome-wide survival study of PDD (Liu et al. 2021), however this was not genome-wide significant in the present study (p=0.016).

We acknowledge several limitations to our study. First, the analysis only included patients of European ancestry and is therefore not generalisable to other populations. In addition, it is possible that some patients were censored as non-demented based on the clinical data we had, but who might have developed dementia if followed-up for a longer period of time. Finally, we grouped patients who developed dementia at any time point together in the design of our study. While our results suggest that the genetic architecture of PDD significantly overlaps with the risk profile of DLB, it is likely that genetic risk factors and associated neuropathology leading to dementia at onset are different from those associated with dementia later in the disease course. We hypothesise that PD patients developing dementia early in the disease course will be genetically closer to DLB, while those developing dementia much later on will present with a different genetic profile. Future studies should aim to identify risk factors leading to a more aggressive disease course in LBD to improve prognosis and care. Finally, further research is needed to clarify if *APOE* is an independent driver of dementia in LBD.

In conclusion, in a pooled analysis of DLB, PD and PDD we have shown that *APOE* and variation at the 5’ end of the *SNCA* gene are the major determinants of LBD with dementia. We have also shown that *LRRK2* G2019S is associated with a dramatically reduced risk of dementia. Although ApoE is associated with dementia, other AD risk loci defined by PRS analysis were not associated with LBD-dementia. Increasing sample sizes in collaborative international studies will help to resolve the disease pathogenesis, the nosological overlap between PDD and DLB and ultimately help to define new treatments.

## Data availability

TPD data is available upon access request from https://www.trackingparkinsons.org.uk/about1/data/. AMP-PD data is available upon registration at https://www.amp-pd.org/. OPDC data is available upon request from the Dementias Platform UK (https://portal.dementiasplatform.uk/Apply). HapMap phase 3 data (HapMap3) is available for download at ftp://ftp.ncbi.nlm.nih.gov/hapmap/. Cis-QTL eQTLGen data was downloaded from (https://www.eqtlgen.org/cis-eqtls.html). eQTL data from eQTL catalogue can be ftp-accessed (https://www.ebi.ac.uk/eqtl/Data_access/). Summary statistics from the PD GWAS (Nalls *et al.*) used to perform the PRS analysis are available from https://pdgenetics.org/resources. The source code is available on GitHub(https://github.com/huw-morris-lab/LBD-case-caseGWAS; https://doi.org/10.5281/zenodo.8335404)

## Acknowledgment

Data used in the preparation of this article were obtained from the AMP-PD Knowledge Platform (https://www.amp-pd.org). AMP-PD is a public-private partnership managed by the FNIH and funded by Celgene, GSK, Michael J. Fox Foundation for Parkinson’s Research, the National Institute of Neurological Disorders and Stroke (NINDS), Pfizer and Verily. TPD and OPDC cohorts are primarily funded and supported by Parkinson’s UK (https://www.parkinsons.org.uk/) and supported by the National Institute for Health and Care Research (NIHR) Clinical Research Network (CRN). The TPD study is also supported by NHS Greater Glasgow and Clyde. The OPDC cohort is also supported by the NIHR Oxford Biomedical Research Centre, based at the Oxford University Hospitals NHS Trust, and the University of Oxford. This research was supported in part by the Intramural Research Program of the National Institutes of Health, National Institute on Aging and National Institute of Neurological Disorders and Stroke.

## Appendix

ORCIDs - Inves⍰ga⍰on of the gene⍰c ae⍰ology of Lewy body diseases with and without demen⍰a (Wu et al.)

Lesley Wu 0000-0001-5464-5603

Raquel Real 0000-0001-8117-742X

Alejandro Mar⍰nez 0000-0001-6191-1703

Ruth Chia 0000-0002-4709-7423

Michael A Lawton 0000-0002-3419-0354

Maryam Shoai: 0000-0003-2499-8533

Catherine Bresner 0000-0003-2673-9762

Leon Hubbard 0000-0002-0064-6755

Cornelis Blauwendraat 0000-0001-9358-8111

Andrew B Singleton 0000-0001-5606-700X

Mina Ryten 0000-0001-9520-6957

Sonja W. Scholz 0000-0002-6623-0429

Bryan J Traynor 0000-0003-0527-2446

Nigel Williams 0000-0003-1177-6931

Michele TM Hu 0000-0001-6382-5841

Yoav Ben-Shlomo 0000-0001-6648-3007

Donald G Grosset 0000-0002-2757-8203

John Hardy: 0000-0002-3122-0423

Huw R. Morris: 0000-0002-5473-3774

