## Supplementary tables and figures for "Investigation of the genetic aetiology of Lewy body diseases with and without dementia"

Supplementary Table 1. Cohort description

| Cohort | Abbreviation | N | Genotyping array |
|---|---|---|---|
| Tracking Parkinson's Disease | TDP | 2000 | Illumina HumanCoreExome array |
| Oxford Parkinson's Disease Centre Discovery Cohort | OPDC | 1082 | Illumina HumanCoreExome-12 v1.1 or Illumina Infinium HumanCoreExome-24 v1.1 |
| BioFIND | BF | 213 | Whole Genome Sequenced |
| Harvard Biomarker Study | HB | 1173 | Whole Genome Sequenced |
| Lewy Body Dementia Study | LB | 4579 | Whole Genome Sequenced |
| LRRK2 Cohort Consortium | LC | 599 | Whole Genome Sequenced |
| Parkinson's Disease Biomarkers Program | PD | 1604 | Whole Genome Sequenced |
| Parkinson's Progression Markers Initiative | PP | 1943 | Whole Genome Sequenced |
| Steady PD - Phase 3 | SY | 329 | Whole Genome Sequenced |
| Sure PD - Phase 3 | SU | 259 | Whole Genome Sequenced |

Supplementary Table 2. Allele frequency

| Groups | | Rs17442721 | Rs34637584 (*LRRK2 G2019S*) |
|---|---|---|---|
| LRRK2 carriers | LBD | 45% | 45% |
| | Non LBD | 47% | 46% |
| Non LRRK2 carriers | LBD | 2% | 0% |
| | Controls* | 2% | 0% |

*Data extracted from non-Finnish Europeans in gnomAD browser (https://gnomad.broadinstitute.org/variant/12-40141971-C-G?dataset=gnomad_r3)

Supplementary Table 3. Dementia in LRRK2 G20219S carriers

| Groups | LBD-D | LBD-ND |
|---|---|---|
| Patients without *LRRK2 G2019S* | 2893 (39%) | 4593 (61%) |
| Patients with *LRRK2 G2019S* | 15 (5%) | 303 (95%) |

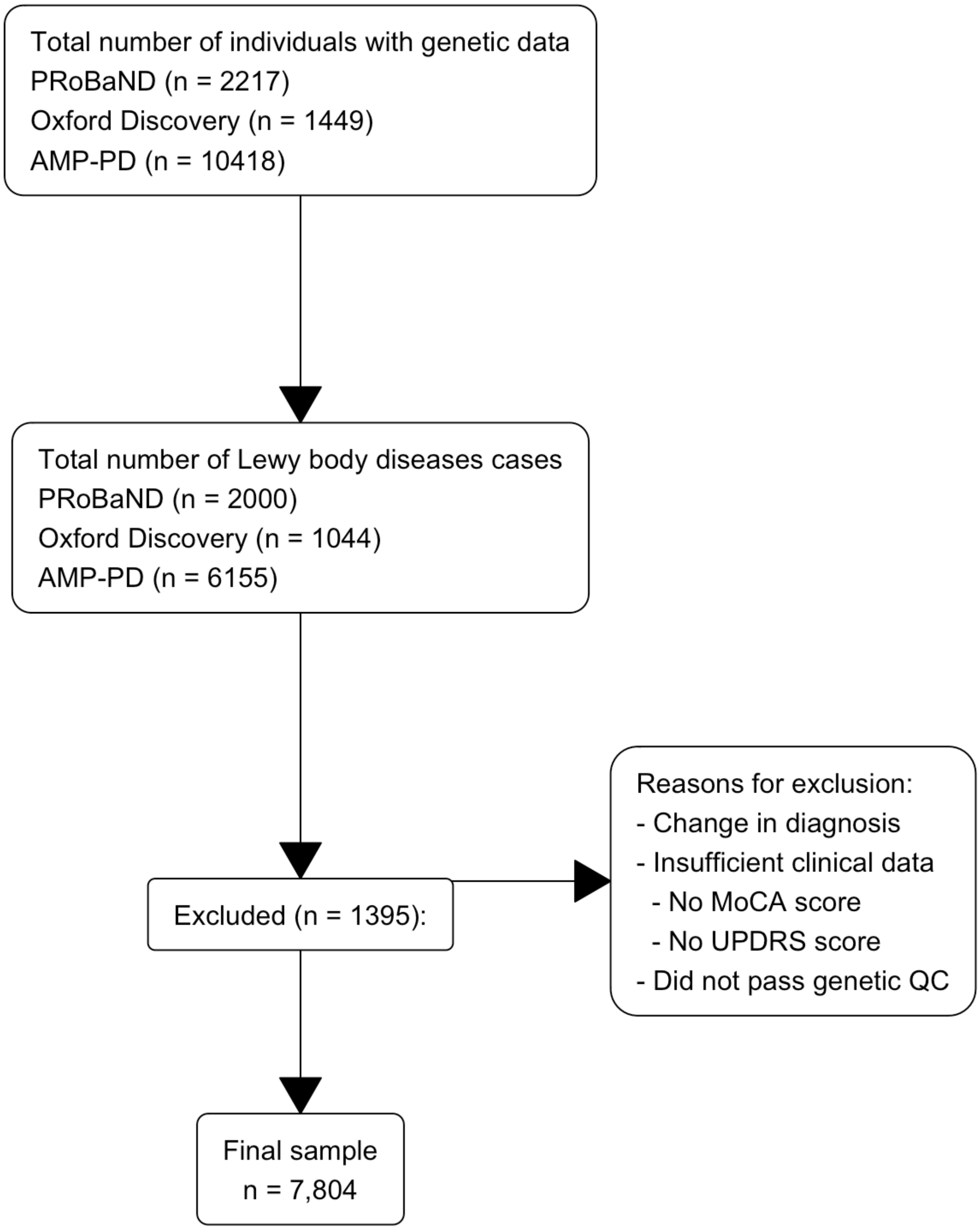


Supplementary figure 1. Sample selection workflow

Samples were selected based on their diagnosis, and whether they passed clinical and genetic data quality control.

1)

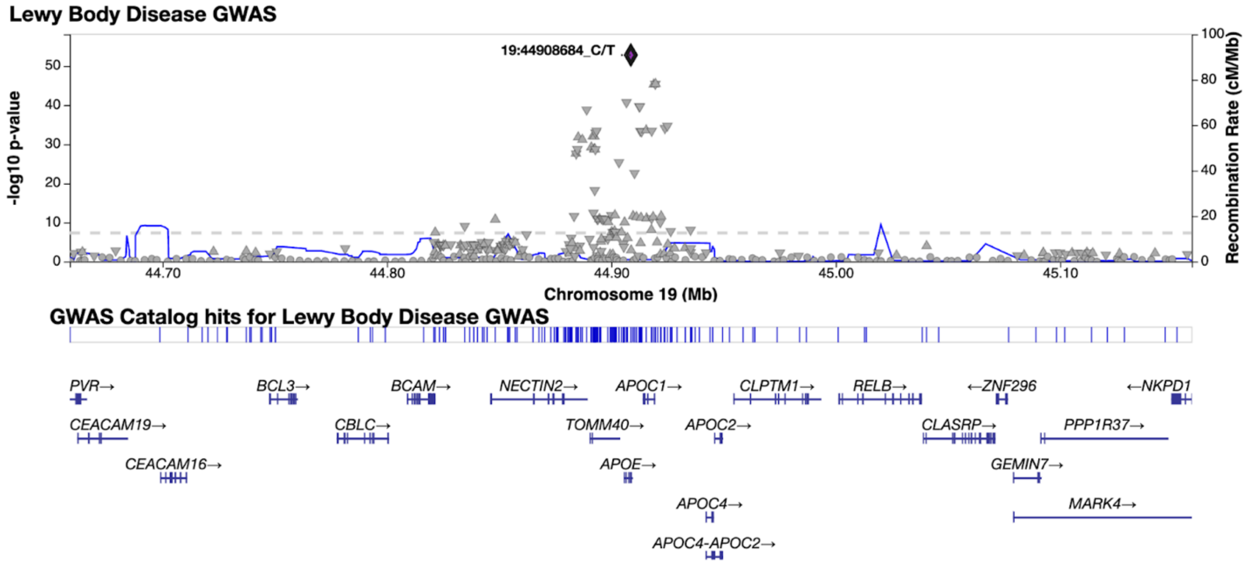
Lewy Body Disease GWAS
19:44908684_C/T
-log10 p-value
Recombination Rate (cM/Mb)
Chromosome 19 (Mb)
GWAS Catalog hits for Lewy Body Disease GWAS
PVR→
BCL3→
BCAM→
NECTIN2→
APOC1→
CLPTM1→
RELB→
←ZNF296
←NKPD1
CEACAM19→
CBLC→
TOMM40→
APOC2→
CLASRP→
PPP1R37→
CEACAM16→
APOE→
GEMIN7→
APOC4→
MARK4→
APOC4-APOC2→

2)

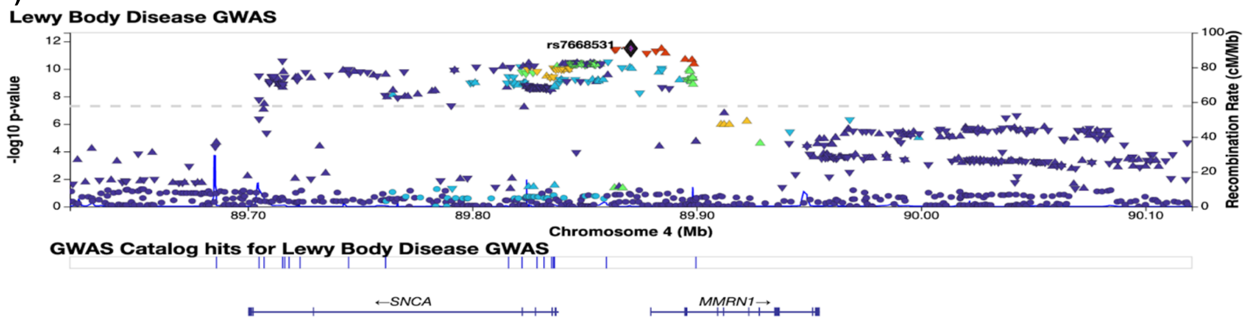
Lewy Body Disease GWAS
rs7668531
-log10 p-value
Recombination Rate (cM/Mb)
Chromosome 4 (Mb)
GWAS Catalog hits for Lewy Body Disease GWAS
←SNCA
MMRN1→

3)

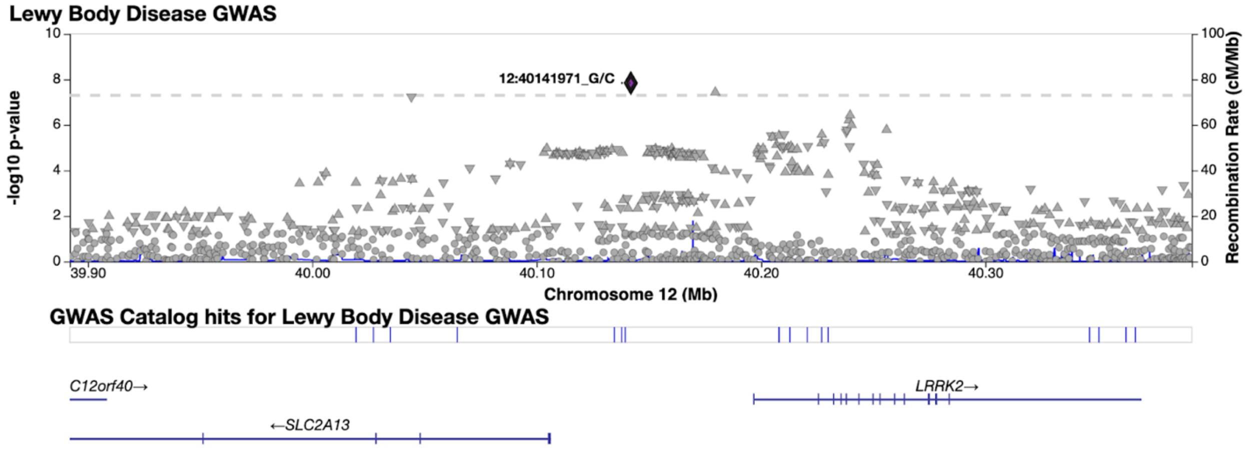


Supplementary Figure 2: Regional association plots of genome-wide significant SNPs

Regional association plots for 1) 19:44908684 (rs429358), 2) 4:89870668 (rs7668531) and 3) 12:40141971 (rs17442721). SNP position and recombination rates are based on GRCh38.

### Appendix

### International LBD Genomics Consortium (iLBDGC)

#### United States

- **Yevgeniya Abramzon, B.Sc.** (Neuromuscular Diseases Research Section, Laboratory of Neurogenetics, National Institute on Aging, Bethesda, MD, 20892, USA)
- **Sarah Ahmed, B.Sc.** (Neurodegenerative Diseases Research Unit, Laboratory of Neurogenetics, National Institute of Neurological Disorders and Stroke, Bethesda, MD, 20892, USA)
- **Camille Alba, Ph.D.** (Department of Anatomy, Physiology and Genetics, Physiology and Genetics, Uniformed Services University of the Health Sciences, Bethesda, MD, 20814, USA)
- **Marilyn S. Albert, Ph.D.** (Department of Neurology, Johns Hopkins University Medical Center, Baltimore, MD, 21287, USA)
- **Dagmar Bacikova, Ph.D.** (Department of Anatomy, Physiology and Genetics, Physiology and Genetics, Uniformed Services University of the Health Sciences, Bethesda, MD, 20814, USA)
- **Matthew J. Barrett, M.D.** (Department of Neurology, University of Virginia School of Medicine, Charlottesville, VA, 22903, USA)
- **Thomas G. Beach, M.D., Ph.D.** (Civin Laboratory for Neuropathology, Banner Sun Health Research Institute, Sun City, AZ, 85006, USA)
- **David A. Bennett, M.D.** (Rush Alzheimer's Disease Center, Rush University, Chicago, IL, 60612, USA)
- **Lilah M. Besser, Ph.D., M.Ph.** (Institute for Human Health and Disease Intervention, Florida Atlantic University, Boca Raton, FL, 33431, USA)

- **Eileen H. Bigio, M.D.** (Mesulam Center for Cognitive Neurology and Alzheimer's Disease, Northwestern University Feinberg School of Medicine, Chicago, IL, 60611, USA)
- **Bradley F. Boeve, M.D.** (Center for Sleep Medicine, Mayo Clinic, Rochester, MN, 55905, USA)
- **Ryan C. Bohannan, B.Sc.** (Department of Neurobiology and Behavior, University of California Irvine, Irvine, CA, 92697, USA)
- **Chad A. Caraway, Ph.D.** (Institute for Memory Impairments and Neurological Disorders, University of California Irvine, Irvine, CA, 92697, USA)
- **Jose-Alberto Palma, M.D., Ph.D.** (Department of Neurology, New York University School of Medicine, New York, NY, 10016, USA)
- **Ruth Chia, Ph.D.** (Neuromuscular Diseases Research Section, Laboratory of Neurogenetics, National Institute on Aging, Bethesda, MD, 20892, USA)
- **Clifton L. Dalgard, Ph.D.** (Department of Anatomy, Physiology and Genetics, Uniformed Services University of the Health Sciences, Bethesda, MD, 20814, USA; The American Genome Center, Collaborative Health Initiative Research Program, Uniformed Services University of the Health Sciences, Bethesda, MD, 20814, USA)
- **Dennis Dickson, M.D.** (Department of Neuroscience, Mayo Clinic, Jacksonville, FL, 32224, USA)
- **Jinhui Ding, Ph.D.** (Computational Biology Core, Laboratory of Neurogenetics, National Institute on Aging, Bethesda, MD, 20892, USA)
- **Kelley Faber, M.Sc.** (Department of Medical and Molecular Genetics, Indiana University School of Medicine, Indianapolis, IN, 46202, USA)
- **Tanis Ferman, Ph.D.** (Department of Psychiatry and Psychology, Mayo Clinic, Jacksonville, FL, 32224, USA)
- **Luigi Ferrucci, M.D., Ph.D.** (Longitudinal Studies Section, National Institute on Aging, Baltimore, MD, 21224, USA)
- **Margaret E. Flanagan, M.D.** (Northwestern University Feinberg School of Medicine, Chicago, IL, 60611, USA)
- **Tatiana M. Foroud, M.D.** (Department of Medical and Molecular Genetics, Indiana University School of Medicine, Indianapolis, IN, 46202, USA)
- **Bernardino Ghetti, M.D.** (Department of Pathology and Laboratory Medicine, Indiana University School of Medicine, Indianapolis, IN, 46202, USA)
- **J. Raphael Gibbs, Ph.D.** (Computational Biology Core, Laboratory of Neurogenetics, National Institute on Aging, Bethesda, MD, 20892, USA)
- **Alison Goate, Ph.D.** (Nash Family Department of Neuroscience, Department of Genetics and Genomic Sciences, and Department of Pathology, Icahn School of Medicine at Mount Sinai, New York, NY, 10029, USA)
- **David Goldstein, M.D.** (Clinical Neurocardiology Section, National Institute of Neurological Disorders and Stroke, Bethesda, MD, 20892, USA)
- **Neill R. Graff-Radford, M.D.** (Department of Neurology, Mayo Clinic Florida, 4500 San Pablo Road South, Jacksonville, FL, 32224, USA)
- **Heng-Chen Hu, Ph.D.** (Department of Anatomy, Physiology and Genetics, Physiology and Genetics, Uniformed Services University of the Health Sciences, Bethesda, MD, 20814, USA)
- **Daniel Hupalo, Ph.D.** (Department of Anatomy, Physiology and Genetics, Physiology and Genetics, Uniformed Services University of the Health Sciences, Bethesda, MD, 20814, USA)
- **Scott M. Kaiser, M.B.A.** (Department of Neuropathology, Indiana University School of Medicine, Indianapolis, IN, 46202, USA)

- **Horacio Kaufmann, M.D.** (Department of Neurology, New York University School of Medicine, New York, NY, 10016, USA)
- **Ronald C. Kim, M.D.** (Department of Neuropathology, School of Medicine, University of California Irvine, Irvine, CA, 92697, USA)
- **Gregory Klein** (Rush Alzheimer's Disease Center, Rush University, Chicago, IL, 60612, USA)
- **Walter Kukull, Ph.D.** (National Alzheimer's Coordinating Center (NACC), University of Washington, Seattle, WA, 98195, USA)
- **Amanda Kuzma, M.Sc.** (Department of Pathology and Laboratory Medicine, Perelman School of Medicine, University of Pennsylvania, Philadelphia, PA, USA)
- **James Leverenz, M.D.** (Cleveland Lou Ruvo Center for Brain Health, Neurological Institute, Cleveland Clinic, Cleveland, OH, 44195, USA)
- **Grisel Lopez, M.D.** (Medical Genetics Branch, National Human Genome Research Institute, Bethesda, MD, 20892, USA)
- **Qinwen Mao, M.D., Ph.D.** (Northwestern University Feinberg School of Medicine, Chicago, IL, 60611, USA)
- **Elisa Martinez-McGrath, Ph.D.** (Department of Anatomy, Physiology and Genetics, Physiology and Genetics, Uniformed Services University of the Health Sciences, Bethesda, MD, 20814, USA)
- **Eliezer Masliah, M.D.** (Molecular Neuropathology Section, Laboratory of Neurogenetics, National Institute on Aging, Bethesda, MD, 20892, USA)
- **Ed Monuki, M.D., Ph.D.** (Department of Pathology & Laboratory Medicine, School of Medicine, University of California Irvine, Irvine, CA, 92697, USA)
- **Kathy L. Newell, M.D.** (Department of Pathology and Laboratory Medicine, University of Kansas Medical Center, Kansas City, KS, 66160, USA)
- **Lucy Norcliffe-Kaufmann, Ph.D.** (Department of Neurology, New York University School of Medicine, New York, NY, 10016, USA)
- **Matthew Perkins, B.Sc.** (Michigan Brain Bank, University of Michigan Medical School, Ann Arbor, MI, 48109, USA)
- **Olga Pletnikova, M.D.** (Department of Pathology [Neuropathology], Johns Hopkins University Medical Center, Baltimore, MD, 21287, USA)
- **Alan E. Renton, Ph.D.** (Department of Neuroscience, Icahn School of Medicine at Mount Sinai, New York, NY, 10029, USA)
- **Susan M. Resnick, M.D.** (Laboratory of Behavioral Neuroscience, National Institute on Aging, Baltimore, MD, 21224, USA)
- **Owen A. Ross, Ph.D.** (Department of Neuroscience & Department of Clinical Genomics, Mayo Clinic Florida, 4500 San Pablo Road South, Jacksonville, FL, 32224, USA)
- **Marya S. Sabir, B.Sc.** (Neurodegenerative Diseases Research Unit, Laboratory of Neurogenetics, National Institute of Neurological Disorders and Stroke, Bethesda, MD, 20892, USA)
- **Clemens R. Scherzer, M.D.** (Precision Neurology Program, Brigham & Women's Hospital, Harvard Medical School, Boston, MA, 02115, USA)
- **Sonja W. Scholz, M.D., Ph.D.** (Neurodegenerative Diseases Research Unit, Laboratory of Neurogenetics, National Institute of Neurological Disorders and Stroke, Bethesda, MD, 20892, USA; Department of Neurology, Johns Hopkins University Medical Center, Baltimore, MD, 21287, USA)
- **Geidy Serrano, Ph.D.** (Civin Laboratory for Neuropathology, Banner Sun Health Research Institute, Sun City, AZ, 85006, USA)
- **Vikram Shakkotai, M.D., Ph.D.** (Department of Neurology, University of Michigan Medical School, Ann Arbor, MI, 48109, USA)

- **Ellen Sidransky, M.D.** (Medical Genetics Branch, National Human Genome Research Institute, Bethesda, MD, 20892, USA)
- **Andrew B. Singleton, Ph.D.** (Molecular Genetics Section, Laboratory of Neurogenetics, National Institute on Aging, Bethesda, MD, 20892, USA)
- **Toshiko Tanaka, Ph.D.** (Longitudinal Studies Section, National Institute on Aging, Baltimore, MD, 21224, USA)
- **Nahid Tayebi, Ph.D.** (Medical Genetics Branch, National Human Genome Research Institute, Bethesda, MD, 20892, USA)
- **Bryan J. Traynor, M.D., Ph.D.** (Neuromuscular Diseases Research Section, Laboratory of Neurogenetics, National Institute on Aging, Bethesda, MD, 20892, USA; Department of Neurology, Johns Hopkins University Medical Center, Baltimore, MD, 21287, USA)
- **Juan C. Troncoso, M.D.** (Department of Pathology [Neuropathology], Johns Hopkins University Medical Center, Baltimore, MD, 21287, USA)
- **Coralie Viollet, Ph.D.** (Department of Anatomy, Physiology and Genetics, Physiology and Genetics, Uniformed Services University of the Health Sciences, Bethesda, MD, 20814, USA)
- **Ronald L. Walton, B.Sc.** (Department of Neuroscience, Mayo Clinic Florida, Jacksonville, FL, 32224, USA)
- **Randy Woltjer, M.D., Ph.D.** (Department of Neurology, Oregon Health & Sciences University, Portland, OR, 97239, USA)
- **Zbigniew K. Wszolek, M.D.** (Department of Neurology, Mayo Clinic Florida, 4500 San Pablo Road South, Jacksonville, FL, 32224, USA)

Canada

- **Sandra E. Black, M.D.** (Institute of Medical Science, Faculty of Medicine, University of Toronto, 1 King’s College Circle, Room 2374, Toronto, ON, M5S 1A8, Canada; Division of Neurology, Department of Medicine, University of Toronto, 27 King's College Circle, Toronto, ON, M5S 1A1, Canada; Heart and Stroke Foundation Canadian Partnership for Stroke Recovery, Sunnybrook Health Sciences Centre, University of Toronto, 1 King’s College Circle, Room 2374, Toronto, ON, M5S 1A8, Canada; Hurvitz Brain Sciences Research Program, Sunnybrook Research Institute, University of Toronto, 2075 Bayview Avenue, Toronto, ON, M4N 3M5, Canada; LC Campbell Cognitive Neurology Research Unit, Sunnybrook Research Institute, University of Toronto, 2075 Bayview Avenue, Toronto, ON, M4N 3M5, Canada)
- **Ziv Gan-Or, M.D., Ph.D.** (Montreal Neurological Institute and Hospital, Department of Neurology & Neurosurgery, McGill University, 3801 University Street, Montreal, QC, H3A 2B4, Canada)
- **Julia Keith, M.D.** (Department of Anatomical Pathology, Sunnybrook Health Sciences Centre, University of Toronto, 1 King’s College Circle, Room 2374, Toronto, ON, M5S 1A8, Canada)
- **Mario Masellis, M.D., Ph.D.** (Cognitive & Movement Disorders Clinic, Sunnybrook Health Sciences Centre, University of Toronto, 1 King’s College Circle, Room 2374, Toronto, ON, M5S 1A8, Canada; Department of Medicine, Division of Neurology, University of Toronto, Toronto, ON, M5S 1A8, Canada; Hurvitz Brain Sciences Research Program, Sunnybrook Research Institute, University of Toronto, Toronto, ON, M5S 1A8, Canada; LC Campbell Cognitive Neurology Research Unit, Sunnybrook Research Institute, University of Toronto, Toronto, ON, M5S 1A8, Canada)
- **Ekaterina Rogaeva, Ph.D.** (Tanz Centre for Research in Neurodegenerative Diseases, University of Toronto, 1 King's College Circle, Room 2111, Toronto, ON, M5S 1A8, Canada)

United Kingdom

- **Dag Aarsland, M.D.** (Department of Old Age Psychiatry, Institute of Psychiatry, Psychology and Neuroscience [IoPPN], King's College London, DeCrespigny Park, London, SE5 8AF, UK)
- **Safa Al-Sarraj, M.D., Ph.D.** (Department of Clinical Neuropathology and London Neurodegenerative Diseases Brain Bank, Institute of Psychiatry, Psychology and Neuroscience [IoPPN], King's College Hospital and King's College London, DeCrespigny Park, London, SE5 8AF, UK)
- **Johannes Attems, M.D.** (Mental Health, Dementia and Neurodegeneration, Translational and Clinical Research Institute, Newcastle University, Newcastle upon Tyne, NE4 5PL, UK)
- **Raffaele Ferrari, Ph.D.** (Department of Molecular Neuroscience, Institute of Neurology, University College London, London, WC1B 5EH, UK)
- **Steve Gentleman, M.D.** (Neuropathology Unit, Division of Brain Sciences, Department of Medicine, Imperial College London, London, W12 0NN, UK)
- **John A. Hardy, Ph.D.** (Department of Neurodegenerative Disease, Reta Lila Weston Laboratories, Queen Square Genomics, UCL Dementia Research Institute, London WC1E 6BT, UK)
- **Angela K. Hodges, Ph.D.** (Department of Old Age Psychiatry, Institute of Psychiatry, Psychology and Neuroscience [IoPPN], King's College London, Maurice Wohl Clinical Neuroscience Institute, London, SE5 9NU, UK)
- **Seth Love, M.D.** (Dementia Research Group, School of Clinical Sciences, University of Bristol, Bristol, BS10 5NB, UK)
- **Ian McKeith, M.D.** (Mental Health, Dementia and Neurodegeneration, Translational and Clinical Research Institute, Newcastle University, Newcastle upon Tyne, NE4 5PL, UK)
- **Christopher M. Morris, Ph.D.** (Mental Health, Dementia and Neurodegeneration, Translational and Clinical Research Institute, Newcastle University, Newcastle upon Tyne, NE4 5PL, UK)
- **Huw R. Morris, M.D., Ph.D.** (Department of Molecular Neuroscience, Institute of Neurology, University College London, London, WC1B 5EH, UK; Department of Clinical and Movement Neuroscience, Royal Free Campus UCL Institute of Neurology, University College London, London, NW3 2PF, UK)
- **Laura Palmer, Ph.D.** (South West Dementia Brain Bank, Bristol Medical School, University of Bristol, Bristol, BS10 5NB, UK)
- **Stuart Pickering-Brown, Ph.D.** (Division of Neuroscience and Experimental Psychology, Faculty of Biology, Medicine and Health, The University of Manchester, Manchester, M13 9PT, UK)
- **Regina H. Reynolds, M.Sc.** (NIHR Great Ormond Street Hospital Biomedical Research Centre, University College London, London, UK; Genetics and Genomic Medicine, Great Ormond Street Institute of Child Health, University College London, London WC1E 6BT, UK; Department of Neurodegenerative Disease, UCL Queen Square Institute of Neurology, University College London, London, UK)
- **Mina Ryten, M.D., Ph.D.** (NIHR Great Ormond Street Hospital Biomedical Research Centre, University College London, London, UK; Genetics and Genomic Medicine, Great Ormond Street Institute of Child Health, University College London, London WC1E 6BT, UK)
- **Alan J. Thomas, M.B.B.S., Ph.D.** (Mental Health, Dementia and Neurodegeneration, Translational and Clinical Research Institute, Newcastle University, Newcastle upon Tyne, NE4 5PL, UK)

- **Bension S. Tilley, M.B.B.S., Ph.D.** (Neuropathology Unit, Division of Brain Sciences, Department of Medicine, Imperial College London, London, W12 0NN, UK)
- **Claire Troakes, Ph.D.** (Department of Old Age Psychiatry, Department of Basic and Clinical Neuroscience, King's College London, DeCrespigny Park, London, SE5 8AF, UK)

Republic of Ireland

- **Francesca Brett, M.D.** (Dublin Brain Bank, Neuropathology Department, Beaumont Hospital, Dublin, D2, Ireland)

France

- **Alexis Brice, M.D.** (Paris Brain Institute, Paris Brain Institute, Sorbonne Universites, 47, boulevard de l'Hôpital, Paris, CS 21 414 – 75646, France)
- **Charles Duyckaerts, M.D.** (Paris Brain Institute, Paris Brain Institute, Sorbonne Universites, 47, boulevard de l'Hôpital, Paris, CS 21 414 – 75646, France)
- **Suzanne Lesage, Ph.D.** (Paris Brain Institute, ICM, Sorbonne Universites, 47, boulevard de l'Hôpital, Paris, CS 21 414 – 75646, France)

Italy

- **Maura Brunetti, M.Sc.** (Rita Levi Montalcini Department of Neuroscience, University of Turin, Turin, 10126, Italy)
- **Andrea Calvo, M.D.** (Rita Levi Montalcini Department of Neuroscience, University of Turin, Turin, 10126, Italy)
- **Antonio Canosa, M.D.** (Rita Levi Montalcini Department of Neuroscience, University of Turin, Turin, 10126, Italy)
- **Adriano Chiò, M.D.** (Rita Levi Montalcini Department of Neuroscience, University of Turin, Turin, 10126, Italy; Institute of Cognitive Sciences and Technologies, C.N.R., Via S. Martino della Battaglia, 44, Rome, 00185, Italy, Azienda Ospedaliero Universitaria Città della Salute e della Scienza, Corso Bramante, 88, Turin, 10126, Italy)
- **Gianluca Floris, M.D.** (Department of Neurology, University Hospital of Cagliari, Via Ospedale, 54, Cagliari, 09124, Italy)
- **Giancarlo Logroscino, M.D., Ph.D.** (Department of Basic Medical Sciences, Neurosciences and Sense Organs, University of Bari "Aldo Moro", Bari, 70121, Italy)
- **Chiara Zecca, B.Sc.** (Department of Basic Medical Sciences, Neurosciences and Sense Organs, University of Bari "Aldo Moro", Bari, 70121, Italy)
- **Jordi Clarimon, Ph.D.** (Universitat Autònoma de Barcelona, Carrer de Sant Quintí, 77-79, Barcelona, 08041, Spain, and Movement Disorders Units, Department of Neurology, University Hospital Mutua de Terrassa, Barcelona, 08221, Spain)
- **Monica Diez-Fairen, M.Sc.** (Memory and Movement Disorders Units, Department of Neurology, University Hospital Mutua de Terrassa, 08221 Barcelona, Spain)
- **Juan Fortea, M.D., Ph.D.** (Universitat Autònoma de Barcelona, Carrer de Sant Quintí, 77-79, Barcelona, 08041, Spain, and Movement Disorders Units, Department of Neurology, University Hospital Mutua de Terrassa, Barcelona, 08221, Spain)
- **Isabel González-Aramburu, M.D., Ph.D.** (Neurology Service, University Hospital Marqués de Valdecilla-IDIVAL-UC, Santander, 39011, Spain)
- **Jon Infante, M.D., Ph.D.** (Neurology Service, University Hospital Marqués de Valdecilla-IDIVAL-UC, Santander, 39011, Spain)
- **Carmen Lage, M.D.** (Neurology Service, University Hospital Marqués de Valdecilla-IDIVAL-UC, Santander, 39011, Spain)

- **Alberto Lleó, M.D., Ph.D.** (Universitat Autònoma de Barcelona, Carrer de Sant Quintí, 77-79, Barcelona, 08041, Spain, and Movement Disorders Units, Department of Neurology, University Hospital Mutua de Terrassa, Barcelona, 08221, Spain)
- **Pau Pastor, M.D., Ph.D.** (Memory and Movement Disorders Units, Department of Neurology, University Hospital Mutua de Terrassa, 08221 Barcelona, Spain)
- **Laura Porcel-Molina, M.D., Ph.D.** (Neurological Tissue Bank, Biobanc-Hospital Clinic - IDIBAPS, C/ Centre Esther Koplowitz, Barcelona, 08036, Spain)
- **Eloy Rodríguez-Rodríguez, M.D., Ph.D.** (Neurology Service, University Hospital Marqués de Valdecilla-IDIVAL-UC, Santander, 39011, Spain)
- **Pascual Sanchez-Juan, M.D., Ph.D.** (Neurology Service, University Hospital Marqués de Valdecilla-IDIVAL-UC, Santander, 39011, Spain)

Luxembourg

- **Rejko Krüger, M.D.** (Luxembourg Center for Systems Biomedicine, University of Luxembourg, Esch-sur-Alzette, Luxembourg, L-4362, Luxembourg; Luxembourg Institute of Health (LIH), University of Luxembourg, Esch-sur-Alzette, Luxembourg, L-4362, Luxembourg; Centre Hospitalier de Luxembourg (CHL), University of Luxembourg, Esch-sur-Alzette, Luxembourg, L-4362, Luxembourg)
- **Patrick May, Ph.D.** (Luxembourg Center for Systems Biomedicine, University of Luxembourg, Esch-sur-Alzette, Luxembourg, L-4362, Luxembourg)

Greece

- **Georgia Xiromerisiou, M.D., Ph.D.** (Department of Neurology, University Hospital of Larissa, University of Thessalia, Mezourlo, Larissa, 41110, Greece)
